# Compendium of clinical variant classification for 2,247 unique *ABCA4* variants to improve genetic medicine access for Stargardt Disease

**DOI:** 10.1101/2023.04.24.23288782

**Authors:** Stéphanie S. Cornelis, Miriam Bauwens, Lonneke Haer-Wigman, Marieke De Bruyne, Madhulatha Pantrangi, Elfride De Baere, Robert B. Hufnagel, Claire-Marie Dhaenens, Frans P.M. Cremers

## Abstract

Biallelic variants in *ABCA4* cause Stargardt disease (STGD1), the most frequent heritable macular disease. Determination of the pathogenicity of variants in *ABCA4* proves to be difficult due to 1) the high number of benign and pathogenic variants in the gene; 2) the presence of complex alleles; 3) the extensive variable expressivity of this disease and 4) reduced penetrance of hypomorphic variants. Therefore, the classification of many variants in *ABCA4* is currently of uncertain significance. Here we complemented the *ABCA4* Leiden Open Variation Database (LOVD) with data from ∼11,000 probands with *ABCA4*-associated inherited retinal diseases from literature up to the end of 2020. We carefully adapted the ACMG/AMP classifications to *ABCA4* and assigned these classifications to all 2,247 unique variants from the *ABCA4* LOVD to increase the knowledge of pathogenicity. In total, 1,247 variants were categorized with a Likely Pathogenic or Pathogenic classification, whereas 194 variants were categorized with a Likely Benign or Benign classification. This uniform and improved structured reclassification, incorporating the largest dataset of *ABCA4*-associated retinopathy cases so far, will improve both the diagnosis as well as genetic counselling for individuals with *ABCA4*-associated retinopathy.

## Introduction

Biallelic variants in *ABCA4* are the cause of Stargardt disease (STGD1) [1], which is the most frequent heritable macular degeneration [2, 3]. It is estimated to affect between 1:6,500 and 1:20,000 people [4-7]. The broad clinical spectrum includes classical STGD1 (onset between 10 and 40 years), cone-rod dystrophy (CRD) (onset before 10 years), and late onset STGD1 (onset after 40 years) [8, 9]. *ABCA4*-associated retinopathy (*ABCA4-*retinopathy) is therefore sometimes used as an umbrella term for all retinal phenotypes associated with *ABCA4*. Due to the recessive nature of the disease, the different levels of severity of variants as well as the large allelic heterogeneity, it is currently challenging to genetically diagnose individuals with *ABCA4-*retinopathy [10].

Furthermore, in determining the pathogenicity of an *ABCA4* variant, multiple other factors should be considered. Several missense and synonymous variants are known to cause splicing defects in *ABCA4* [11-13], therefore missense variants at positions that are not conserved and synonymous variants cannot simply be dismissed as likely benign. In addition, reduced penetrance has been reported for multiple hypomorphic variants in *ABCA4* [14, 15] meaning that variants with a relatively high allele frequency can still be pathogenic. One might expect that when two *ABCA4* variants are detected in a person with a STGD1-like phenotype, these must be biallelic and causal. However, it is not uncommon to find variants to be *in cis* in *ABCA4* and multiple complex alleles have been described [16-18]. Several studies indicate that in cases with a single variant in *ABCA4* or a single complex *ABCA4* allele, the disease-causing variant(s) can be found in other genes [19-21]. Moreover, the effect of severity on protein function varies enormously among *ABCA4* pathogenic variants. Two null alleles (alleles with a complete loss of function) can cause legal blindness before the age of 10 [22-25], whereas mild variants usually only cause disease when present *in trans* with a null or severe allele, often with a late age at onset, which also show reduced penetrance [15]. The most extreme mild variant is c.5603A>T, which only seems to cause visual impairment in ∼5% of individuals when *in trans* with a deleterious allele [14, 26]. This further illustrates the complexity of *ABCA4*-retinopathy and the complexity that needs to be dealt with when classifying *ABCA4* variants.

Consequently, many individuals with *ABCA4*-retinopathy are currently not genetically diagnosed, as only one pathogenic variant or allele or biallelic variants of uncertain significance have been identified. For these individuals, it is important to know whether the variants they have are pathogenic. Currently, the pathogenicity of *ABCA4* variants is critical as many gene therapy trials are under investigation, for which only individuals with biallelic pathogenic alleles are eligible. Moreover, as the carrier rate of pathogenic *ABCA4* variants in the general population is relatively high [6, 27], carrier analysis is performed frequently to determine the risk for future offspring and in these cases, it is important to know whether identified variants are pathogenic.

Databases such as the *ABCA4*-Leiden Open (source) Variation Database (LOVD) (https://databases.lovd.nl/shared/genes/ABCA4) and ClinVar (http://www.clinvar.com/), provide a wealth of information for *ABCA4* variants including pathogenicity classifications. However, there is discordance between databases. The *ABCA4*-LOVD reports that up to 85% of the variants could be pathogenic, whereas in ClinVar, ∼40% of the variants are reported to be likely pathogenic, which might be because many novel variants of uncertain significance are reported in ClinVar. Both the *ABCA4*-LOVD and ClinVar also allow for variable classifications as they are submitter-reported. Therefore, due to the variety of classification methods, it is difficult to truly assess and compare the pathogenicity of different variants.

A clear and broadly used pathogenicity classification system is the ACMG/AMP classification as described by Richards et al. in 2015 [28]. This classification method incorporates information such as type of variant, *cis/trans* criteria, variant frequency, phenotype, functional studies, segregation and *in silico* predictions. All information can be collected per variant and can easily be combined to a final pathogenicity classification with five tiers: Benign, Likely benign, Variant of Uncertain Significance (VUS), Likely Pathogenic or Pathogenic. Since it is consistently used worldwide, it allows easy interpretation and comparison of pathogenicity levels.

In order to increase the knowledge on the pathogenicity of genetic variants in *ABCA4*, we collected all the published data on *ABCA4*-retinopathy cases that have been published up to 31 December 2020 and uploaded them into the *ABCA4*-LOVD. Here, we adapted ACMG/AMP classifications specifically for *ABCA4* and applied them to all 2,247 variants present in the *ABCA4*-LOVD.

## Methods

We collected all papers published until 31 December 2020, which contain likely pathogenic *ABCA4* variants in individuals with autosomal recessive retinopathy by searching the following search terms in PubMed:

(ABCA4[All Fields] OR ((“Stargardt disease”[All Fields] OR “Macula Lutea”[All Fields]) AND (“Genetics”[All Fields] OR “mutation”[All Fields] OR “Sequence Analysis”[All Fields] OR “gene panel”[TiAb]))) OR (“Retinal Dystrophies”[All Fields] AND (“mutation”[All Fields] OR “Sequence Analysis”[All Fields] AND “gene panel”[TiAb]))

Reported variants were collected per patient as well as additional available data such as gender, type of vision impairment, ethnicity, geographical origin, age at onset, phenotype at onset, segregation data, consanguinity status and other remarks. The data were supplemented with data from 412 persons with *ABCA4*-retinopathy from PreventionGenetics (a division of Exact Sciences). All data have been uploaded into the *ABCA4*-LOVD (www.lovd.nl/ABCA4).

### *ABCA4*-LOVD

All *ABCA4* variant data from the *ABCA4*-LOVD were downloaded on 5 April 2022.

### Nomenclature

The annotation of all variants was done according to Human Genome Variation Society (HGVS) nomenclature guidelines where possible and is based on the GRCh37 hg19 genomic coordinates, gene location NM_000350.3. All variants were checked using the Batch Validator of the online VariantValidator tool [29].

### ACMG/AMP classification

All variants were classified according to ACMG/AMP variant classification guidelines described by Richards et al., 2015, the updated recommendations from ClinGen reported at https://www.clinicalgenome.org/working-groups/sequence-variant-interpretation/ and the naturally scaled ACMG/AMP point system of Tavtigian et al. 2020 [30]. Our project group, consisting of experts on *ABCA4* genetics and ophthalmology, decided how best to apply the ACMG/AMP categories and ClinGen recommendations to *ABCA4* variants, which is summarized in Figure 1 and can be found in more detail in the Supplemental Materials & Methods.

**Figure 1.**
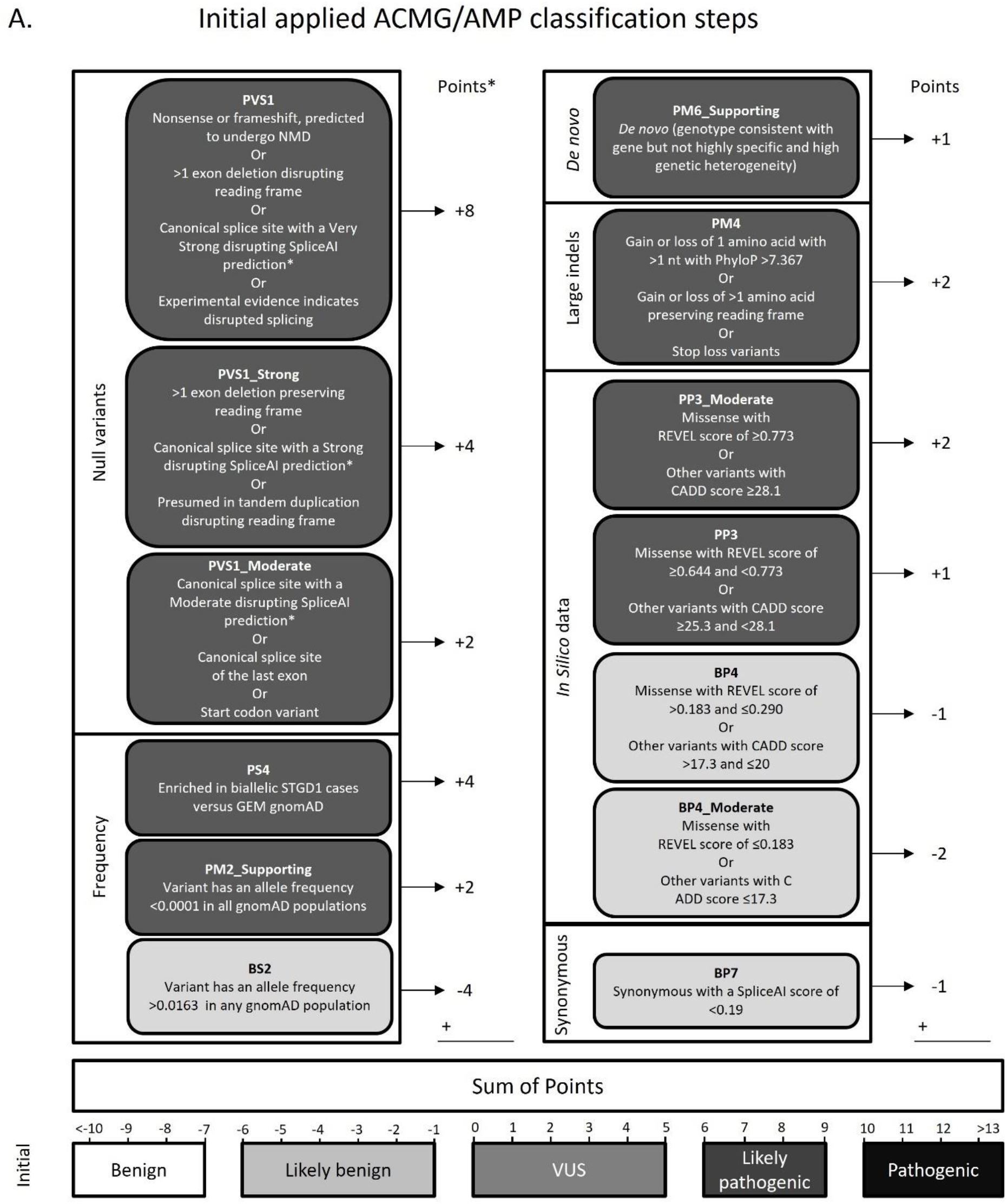
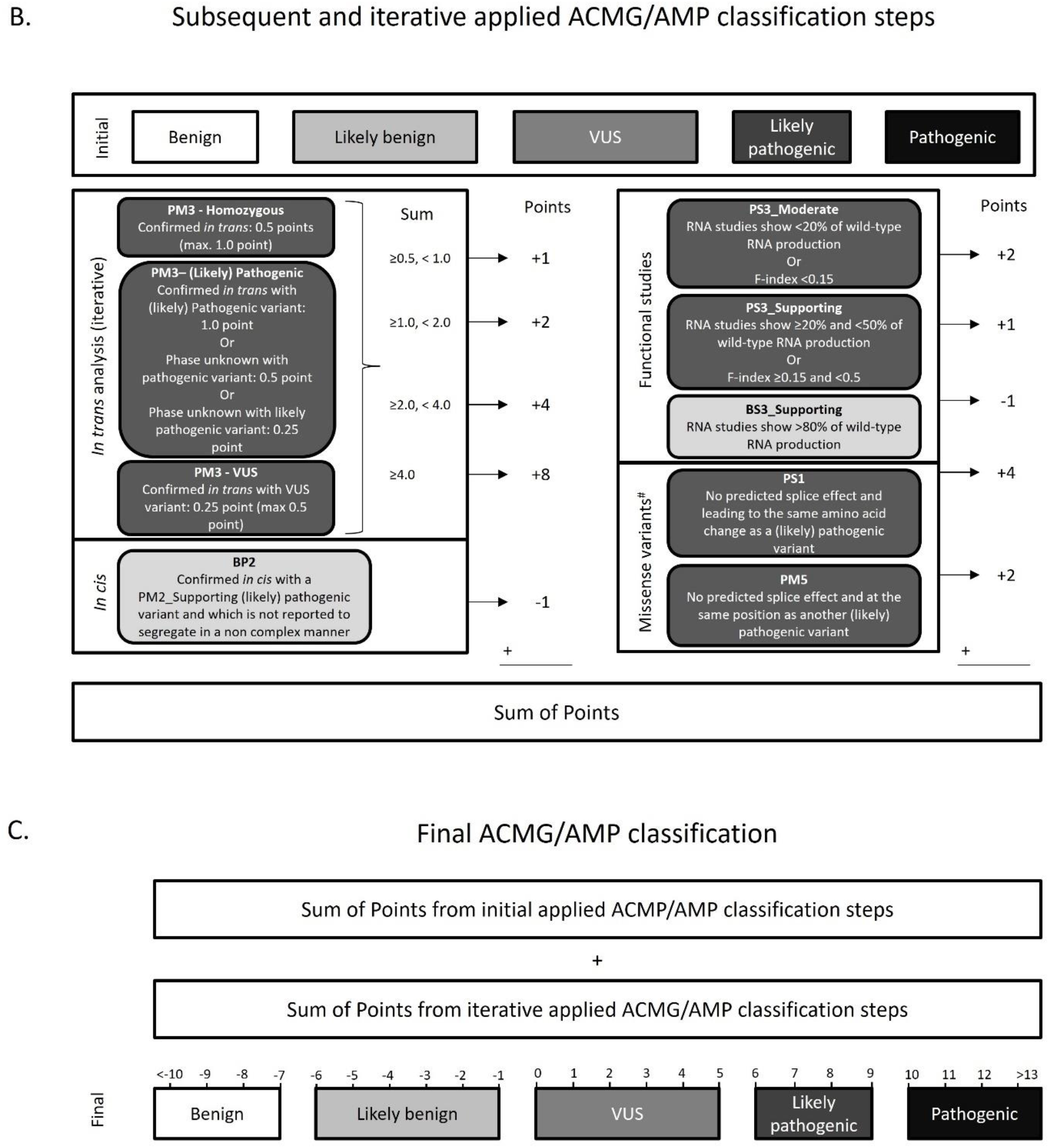
Applied ACMG/AMP classification steps. A. Initial steps to classify *ABCA4* variants from LOVD based on ACMG/AMP classification. For each step, a number of points is awarded and their sum is used to determine the initial ACMG/AMP classification according to Tavtigian et al., 2020 [30]. GEM gnomAD data are described in Cornelis et al., 2022 [27]. B. Subsequent and iterative classification steps based on the initial ACMG/AMP classification. #The missense classification steps were not iterated to avoid circular reasoning. C. Sum of points from initial steps and subsequent and iterative steps are combined into a final ACMP/AMP classification score.

## Results

### Published *ABCA4*-retinopathy cohort

After the removal of likely duplicates, the collection of data contained variants from 10,391 likely *ABCA4-*retinopathy individuals, of which 3,411 were already reported in our 2017 study [31]. The cohort contained 6,240 likely biallelic cases and 4,151 mono-allelic cases. Among these were 943 non-consanguineous homozygous cases and 127 consanguineous homozygous cases. A total of 2,095 unique variants were identified in the cohort. All data were uploaded to the *ABCA4*-LOVD database www.lovd.nl/ABCA4.

### ACMG/AMP classification

ACMG/AMP classifications were given to all variants from the published cohort as well as other *ABCA4* variants from the *ABCA4* LOVD (Table S1) and were annotated with the predicted pathogenicity severity according to Cornelis et al. [27] when available. Results of each ACMG/AMP classification step can be found in Tables S2-S10. After applying the non-iterative classification steps, 1,276 variants could be categorized as Benign (n=8), Likely Benign (n=191), Likely Pathogenic (n=498) or Pathogenic (n=579), and 973 variants as VUS. The iterative classification steps changed the number of variants categorized as (Likely) Benign or (Likely) Pathogenic to 1,441 (Table 1 and Figure 2A). Of note, at the end of the analysis, 49 null variants reach ‘Pathogenic’ without the PVS1 criterium and >10% of variants associated with *ABCA4-*retinopathy are loss of function confirming that the use of PVS1 flowchart is correct.

**Table 1.** Overview of the final ACMG/AMP categorization of *ABCA4* variants from LOVD

| Benign | Likely Benign | VUS | Likely Pathogenic | Pathogenic |
| --- | --- | --- | --- | --- |
| 12 | 182 | 806 | 451 | 796 |

**Figure 2.**
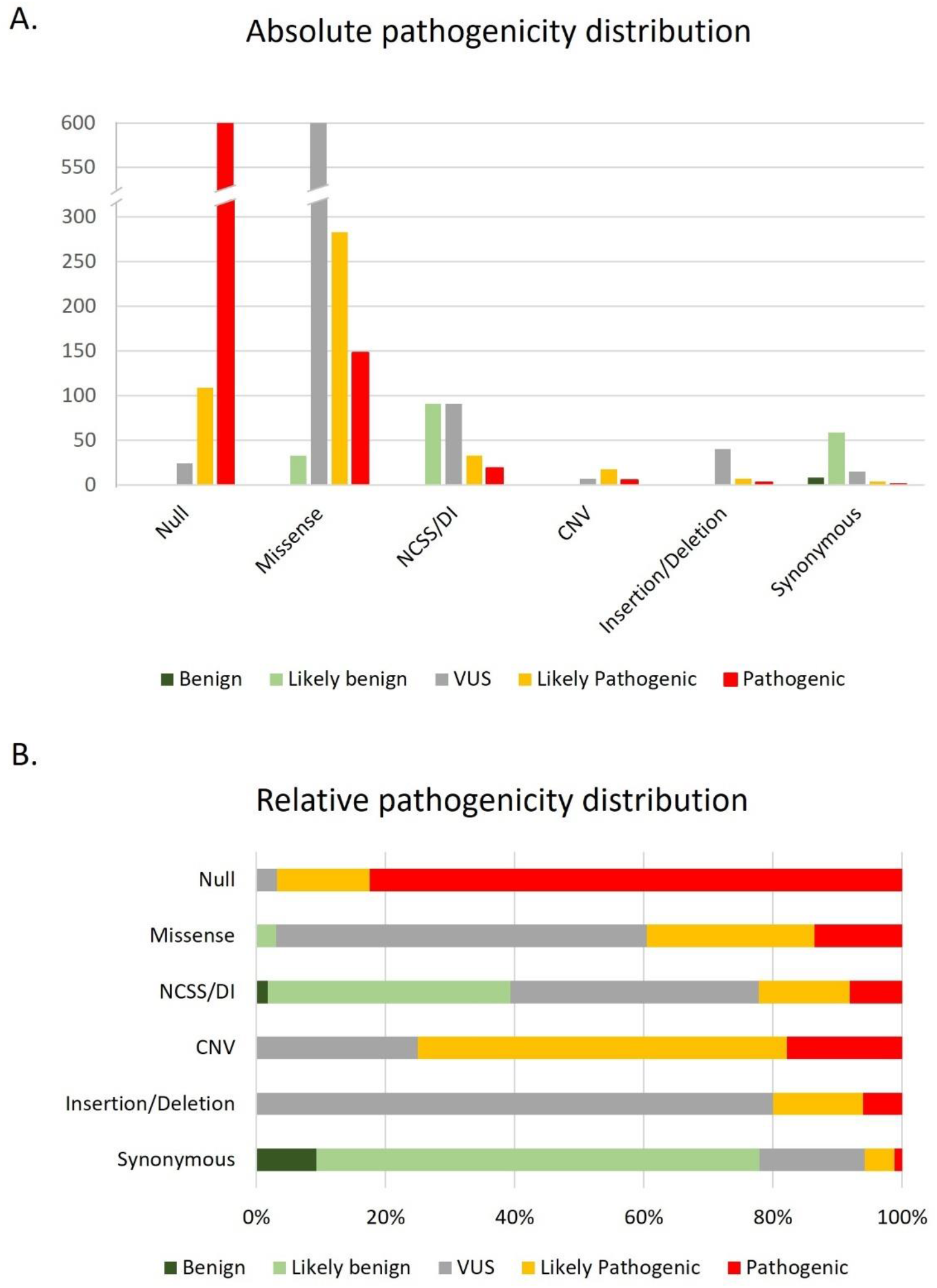
Distribution of ACMG/AMP classifications among different types of variants (null, missense, noncanonical splice site [NCSS]/deep intronic [DI], copy number variation [CNV], [in-frame] insertion/deletion and synonymous). A. Absolute occurrence of different pathogenicity classifications per variant type. B. Relative pathogenicity classifications per variant type.

### Null variants

In total, 752 null variants were reported. Interestingly, 24 of these variants are still classified as VUS after the application of the ACMG/AMP classification system (Figure 2). Compared to other null alleles, the splice predictions for these 24 variants indicated that alternative in-frame splicing could occur or in-frame deletions were predicted. Furthermore, their occurrence in the dataset was too low to reach significance in the frequency analysis.

### Missense variants

A group of variants that is very difficult to interpret without an ACMG/AMP classification are missense variants. Here, we were able to classify 431 missense variants as (Likely) Pathogenic and 33 missense variants as Likely Benign. At the end of the analyses, 627 variants were classified as VUS (Figure 2).

### Synonymous variants

Of the 86 synonymous variants, five variants could be classified as (Likely) Pathogenic and 67 variants as (Likely) Benign (Figure 2).

### Frequency analysis

In the frequency analysis, 856 variants reached significant enrichment in the likely biallelic dataset compared to the GAM gnomAD control dataset after correction for multiple testing with the Benjamini Hochberg method [32]. Interestingly, two variants, c.5603A>T and c.4253+43G>A, were significantly enriched, but had an odds ratio close to 1 (1.10 and 1.49 respectively) without having 1 in the confidence interval. This is a smaller odds ratio than 3, which is generally considered by Richards et al. [28] as the minimal odds ratio for variants with a modest Mendelian effect size. This effect is likely due to the reduced penetrance of these variants. Furthermore, 1,125 variants were not enriched in the likely biallelic dataset but had an allele frequency below 0.0001 in all gnomAD populations and therefore got ‘PM2_Supporting’ evidence.

### *In silico* analyses

SpliceAI scores were given to all variants <50 nucleotides apart from indels. Where possible, indels were given a CI-SpliceAI score. In total, 472 variants had a (CI-)SpliceAI score >0.2 and received PP3 as minimal evidence. In parallel, REVEL and CADD scores were given to missense variants and other variants, respectively. In total, 1,141 variants received ‘PP3_Moderate’ evidence, of which 658 variants were missense variants. On the lower end of the spectrum, 194 variants received ‘BP4_Moderate’. Interestingly, of those 194 variants, only 6 were missense variants and 80 were other exonic variants. Finally, 180 additional variants received PP3, and 48 variants received BP4. Of note, when comparing CADD and REVEL scores for missense variants, it was noticed that only 7 variants received ‘BP4_Moderate’ based on REVEL, while 124 variants would have received ‘BP4_Moderate’ if the CADD score would have been used (Figure S1).

### Segregating complex alleles

The dataset contained a total of 65 unique alleles containing two or more variants (also known as complex alleles) for which segregation had been reported (Table S13). Of note, many individuals with multiple *ABCA4* variants were not reported to have undergone segregation analysis, resulting in a relatively low number of known complex alleles. The three most frequently reported complex alleles were c.[1622T>C;3113C>T], c.[5461-10T>C;5603A>T] and c.[2588G>C;5603A>T]. Forty-three complex alleles were reported only once. Furthermore, twelve complex variants, including the three most frequent complex variants mentioned above, reached a Likely Pathogenic categorization based solely on the *in trans* analysis without correction for allele frequency. The nine other Likely Pathogenic complex allele variants were c.[302+68C>T;4539+2028C>T], c.[769-784C>T;5603A>T], c.[983A>T;3106G>A], c.[1715G>A;2588G>C], c.[3758C>T;5882G>A], c.[4222T>C;4918C>T], c.[4253+43G>A;6006-609T>A], c.[4469G>A;5603A>T] and c.[4926C>G;5044_5058del]. Of note, the pathogenicity scores of alleles with two variants do not have to reflect that both single variants have the same scores. Therefore, for reference, ACMG/AMP classifications of the single variants contained in the complex allele are given in Table S13 as well.

### Frequent pathogenic variants

Two previously known pathogenic variants met the BS1 criterium – an allele frequency of >0.0163 in any gnomAD population - while also reaching a (Likely) Pathogenic classification. The c.5882G>A variant has an allele frequency of 0.023 in the gnomAD Ashkenazi Jewish population. The other variant is c.6320G>A, which has an allele frequency of 0.021 in the gnomAD African population. For each gnomAD population, the three most frequent (Likely) Pathogenic variants are reported in Table S11. Interestingly, three relatively less well-known variants, c.2791G>A, c.2966T>C, and c.2971G>C have a very high frequency (0.0030-0.0073) in the gnomAD African population, while they are less frequent in other populations. Given their high allele frequency, it will be of interest to investigate if they show reduced penetrance. Previously reported frequent pathogenic variants and their updated ACMG/AMP classification can be found in Table S12. Interestingly, the variant c.2588G>C, earlier described as ‘North European’, has a high frequency in the gnomAD South Asian population as well.

## Discussion

In this study, we were able to classify 1,441 of 2,247 *ABCA4* variants with an ACMG/AMP classification other than VUS, based on the point system of Tavtigian et al. [30], which is more than twice as much as our 2017 classification [31]. In total, 1,247 variants were classified as either Likely Pathogenic or Pathogenic, while 194 variants were classified as either Likely Benign or Benign. This will be of important value to clinical geneticists, individuals affected by *ABCA4*-retinopathy and their family members, and ongoing clinical trials for gene-specific therapies.

The ACMG/AMP classification is designed in a way that Likely Pathogenic and Likely Benign variants are classified with ≥90% certainty. Although this is usually interpreted as a very reliable classification, it should be mentioned that when a large group of variants is classified, some variants will receive an incorrect classification. Furthermore, it is important to mention that the binary pathogenicity framework that the ACMG/AMP classification system is based on, currently categorizes a variant as pathogenic if it can cause disease, even if it does not always cause disease *in trans* with another pathogenic variant: In *ABCA4*, multiple variants have been reported to show reduced penetrance [15], meaning that they are not always disease causing *in trans* with another pathogenic variant. Here, we annotated variants that have been reported to show reduced penetrance (Table S1). Reduced penetrance is reflected in both discordance in families with *ABCA4*-retinopathy as well as in a low odds ratio in enrichment studies. Interestingly, the well-known variant c.5603A>T, which shows a very low penetrance when *in trans* with a severe or null allele (5%) [14] is classified as a VUS. It should be noted that in the collection of variant data, it was clear that c.5603A>T was underreported in the data from before 2017, since it has recently been found to co-occur *in cis* with c.2588G>C in individuals with *ABCA4*-retinopathy. Most c.2588G>C occurrences in literature, however, do not mention a co-occurrence with c.5603A>T. This means that the pathogenicity of c.2588G>C may be overclassified here, while that of c.5603A>T may be underclassified, especially since the pathogenicity of c.2588G>C as the only variant in an allele is questionable based on allele frequency calculations in a large cohort [26]. Similarly, the mild reduced penetrant variants c.4253+43G>A and c.769-784C>T, which show a splice defect *in vitro* [16], are classified as VUS and as likely benign, respectively. A possible explanation for this is that deep intronic variants – other than the ones reported by Braun et al. in 2013 [33] - are underreported in literature, mainly because they were missed in most studies that used targeted *ABCA4* sequencing or WES and additionally because their interpretation can be challenging. Therefore, they are less likely to reach significant enrichment in the dataset and may be excluded from the PM3 criterium *in trans* classification when they are not reported. This illustrates the difficulty of classifying frequent mild variants that show reduced penetrance. Therefore, relatively frequent variants occurring in patients harboring a severe variant *in trans* should be treated with caution and larger studies are necessary to identify additional reduced penetrance variants in *ABCA4*.

Modifiers seem to play a role in *ABCA4*-retinopathies and may explain this reduced penetrance of variants. A sex imbalance has been reported for individuals having mild likely reduced penetrant variants [15] and common *PRPH2* variants and rare *ROM1* variants have been reported to act as modifiers of *ABCA4*-retinopathies [34]. In three Dutch families with biallelic sibling pairs carrying c.5603A>T *in trans* with another severe *ABCA4* variant, not all siblings were affected by *ABCA4*-retinopathy [14]. Kjelström and others may also have found two unaffected male individuals over 50 that had both c.5603A>T and a severe variant [35]. Modifiers may similarly explain the reported high variety in disease course of *ABCA4*-retinopathy [36]. Furthermore, modifiers might also aggravate *ABCA4-*retinopathies of individuals: Leber congenital amaurosis, causing severe vision loss in the first year of life, is usually not associated with variants in *ABCA4*, but Panneman and others identified probands in which two *ABCA4* null alleles are hypothesized to cause Leber congenital amaurosis [37] and (Panneman, Koenekoop, Cremers, unpublished data).

Another important point to raise for recessive disease is that variants leading to a protein with reduced but not abolished expression and/or function may not always be disease-causing, depending on the variant *in trans*. For example, if a variant reduces protein expression to 45% compared to WT, then its occurrence next to a null variant is likely disease causing. A homozygous occurrence of this variant, however, will lead to expression only just below that of an individual with a null allele in addition to a WT allele. This may cause a situation similar to a combination of a hypomorphic variant next to a null allele, where modifiers may determine whether an individual will be affected or not. In other words, it is possible that all *ABCA4* variants are on a spectrum based on residual protein function and resulting cellular dysfunction and that the combined severity of *ABCA4* variants and modifiers determine disease penetrance and severity.

Furthermore, several studies indicate that pathogenic variants in other genes can be responsible for *ABCA4-*retinopathy even when one Likely Pathogenic *ABCA4* variant is present in the patient. Disease causing variants can sometimes be found in genes like *PRPH2* and *PROM1*, but also in less common genes associated with *ABCA4*-like disease like *BEST1, CDHR1, CERKL, CNGA3, CRX, ROM1, RPE65* [19, 21, 38].

There are some limitations to this study. The first one is associated with PS4, the allele frequency analysis. First, the GEM BAP control dataset used is based on the reported ethnicity, which may not correspond with the gnomAD population that they were matched with, since ethnicity is a social construct and gnomAD populations are for a big part based on principal component analyses [39] and it is unknown to what extent those overlap. Second, for those patients without reported ethnicity, the GEM BAP gnomAD has incorporated estimated ethnicity based on population statistics. However, there may be a bias in the ethnicity of patients who are able to either afford healthcare, who have the option to take part in a study, or who feel safe to take part in a study since, for example, historic transgressions have been made against Black research participants [40]. Finally, as labs that study *ABCA4*-retinopathy and report individuals with *ABCA4-*retinopathy are unequally distributed over the world, a bias in the genetics of subpopulations of those regions could occur. Therefore, it is likely that, to some degree, population stratification will have affected the results of the allele frequency test. Currently, both genetic and disease data of white individuals with *ABCA4-*retinopathy are overrepresented, creating an imbalance in understanding of the genetic cause of *ABCA4*-retinopathy and treatment options between these individuals and individuals of color with *ABCA4*-retinopathy. In order to improve healthcare for everyone, it is therefore important that rare genetic variants in individuals with under reported genetic ancestry in literature are studied more to improve the knowledge on all genetic variants, and that variants with a high frequency difference between populations are investigated more closely to study their effect since differences in identified variants and numbers of *ABCA4-*retinopathy cases between different ethnicities have been reported [41].

Moreover, based on the final classification, 298 total variants from the biallelic dataset (2.3%) are likely benign or benign. This means that up to 298 cases from this dataset are not actually known biallelic, which might indicate that those cases are not actually *ABCA4*-retinopathy cases, which could create a bias in the enrichment analysis for the variants present *in trans*.

Another limitation of our approach is the underlying variant detection techniques used to identify variants. Especially older data may lack reporting of deep intronic variants that might either be missed as single alleles or *in cis* with reported variants. This underrepresentation of deep intronic variants may cause them to go unnoticed in the PS4 step. Furthermore, more comprehensive *ABCA4* testing strategies are used in countries with good access to genetic testing, which means that new variants and deep intronic variants may be underreported in datasets from countries where access to these techniques is limited. Therefore, the absence of the PS4 category should not be interpreted as proof of a variant not being enriched in the dataset.

Furthermore, the ACMG/AMP guidelines and recommendations warn for the use of functional studies. We indeed encountered that a variant, c.4539+2028C>T, which likely is a pathogenic variant based on genotype-phenotype correlations [33], shows a higher percentage of WT RNA in patient-derived retinal-like cells than expected [42]. We decided to remove this data point from the dataset as an outlier in the classification based on functional studies in step BS3. In addition, other variants, e.g., c.4539+2001G>A and c.1937+435C>G, show a relatively high percentage of WT RNA production, 75% and 55% respectively, in patient-derived retinal-like cells [42] and midigene assays respectively [16], while genotype-phenotype correlations show that these variants are likely pathogenic [33, 43]. This indicates that other variants could show a similar pattern, meaning that intronic variants causing a relatively high amount of WT RNA, may nevertheless be pathogenic. Therefore, results from midigene assays and patient-derived retinal-like cells should be interpreted with caution. However, since most variant results seem to correlate with their pathogenicity, BS3_Supporting was deemed to be of proper evidence strength.

In addition, in the use of *in silico* predictions, the CADD score was used for non-missense variants. However, the applied cut-offs were based on the study of Pejaver and others [44] where they were based on missense variants only. When comparing REVEL and CADD scores for *ABCA4* variants, CADD scores seem to lead to a more benign category. Therefore, we decided to increase the range of scores leading to no *in silico* score from 22.7-25.3 to 20-25.3 to avoid incorrectly classifying noncanonical and synonymous variants as benign since 20 is often used as a cut-off between a benign and a pathogenic indication.

Finally, the *ABCA4* variant data set analyzed here mostly stems from 421 peer-reviewed publications as well as data from PreventionGenetics (a division of Exact Sciences). In the future, it would be very valuable to also include variant data collected in many academic and non-academic diagnostic centers worldwide. This will be challenging as privacy rules may prevent data sharing and different privacy rules in different countries likely create a bias in the data. Furthermore, data currently existing in different online databases may show overlap and are not all curated.

With the advent of novel therapies, it is essential to have an accurate genetic diagnosis, which emphasizes the importance of the classification of variants and proper guidelines. *ABCA4* variant classification is challenging due to the higher variant frequency, presence of complex alleles and hypomorphic variants with reduced penetrance. The adapted ACMG/AMP classifications provided in this study, in combination with the earlier established severity assessments for *ABCA4* variants, will facilitate the interpretation of diagnostic results for *ABCA4*-retinopathies, the most common recessive retinal disease.

## Supporting information

Table S1 - ACMG Categorisation

Table S2 - PVS1

Table S13 - Complex alleles

Table S11 - Frequent pathogenic variants

Table S12 - Previously published frequent pathogenic variants

VariantValidator results

## Data Availability

The collected data from affected individuals from literature used to support the findings of this study have been deposited in the ABCA4-LOVD: www.lovd.nl/ABCA4. Additional data produced in the present study are available upon reasonable request.

## Acknowledgements

We would like to acknowledge Kerry Goetz for sharing the *ABCA4* variant data of patients from the eyeGENE network [45] and Bob Molday for sharing F-index data and his advice on interpretation of the F-index.

## Author contributions

Conceptualizations: S.S.C., F.P.M.C. Data collection: S.S.C. Data sharing PreventionGenetics: M.P. Methodology: S.S.C., M.B., L.H-W., M.D.B., E.D.B., R.B.H., C-M.D., F.P.M.C. Data analysis: S.C. Data classification: S.C. Supervision: F.P.M.C. Writing - original draft: S.C., C-M.D., F.C. Writing - reviewing & editing: S.S.C., M.B., L.H-W., M.D.B., M.P., E.D.B., R.B.H., C-M.D., F.P.M.C.

## Supplementary Material

A file containing supplementary material pertinent to this manuscript is available. This supplementary information consists of 13 tables, 2 figures, and detailed methods, as follows: Table S1: All variants with ACMG/AMP classifications (column H) and severity subclassifications from [27] (column I). ACMG/AMP classifications are based on the point system as described by Tavtigian and others [30]. In short, Supporting, Moderate, Strong and Very Strong evidence is combined into a score where each type of evidence gives a score of 1, 2, 4 or 8 respectively, where Pathogenic evidence gives a positive score, and Benign evidence gives a negative score. The resulting total score per variant results in a Benign (<-6), Likely Benign (−1 – -6), VUS (0 – 5), Likely Pathogenic (6 – 9) or Pathogenic (>9) classification. Table S2: ACMG/AMP classification step PVS1 Null variants. Table S3: ACMG/AMP classification step PM6 *De novo* variants. Table S4: ACMG/AMP classification step PS4 Variant Frequency and Use of Control Populations. Table S5: ACMG/AMP classification step PM4 Protein length changes due to in-frame deletions/insertions and stop losses. Table S6: ACMG/AMP classification steps PP3 and BP4 Computational (*in silico*) data. Table S7: ACMG/AMP classification step BP7 Synonymous variants. Table S8: ACMG/AMP classification steps BS1 and PM2 Variant Frequency and Use of Control Populations. Table S9: ACMG/AMP classification steps PS1 and PM5 Same amino acid change and Novel missense at the same position. Table S10: ACMG/AMP classification steps PS3 and BS3 Functional studies. Table S11: Three most frequent (Likely) Pathogenic variants per gnomAD population. Table S12: Previously reported frequent Pathogenic variants based on literature. Table S13: Published segregating complex alleles. Figure S1: *In silico* comparison of CADD and REVEL for missense variants in *ABCA4. In silico* comparison of *ABCA4* missense variants. CADD PHRED values are plotted against REVEL values. Cut-off values between ‘BP4_Moderate’ and ‘BP4’, and between ‘PP3’ and ‘PP3_Moderate’ are shown in green and red respectively, vertically for CADD scores and horizontally for REVEL scores. Overall, it can be observed that the variants reach a higher category for REVEL than for CADD. Figure S2: Correlation between variant F-indexes and age of onset in patients. Correlations between *ABCA4* variants’ F-index and the corresponding log 10 and square root transformed age at onset for homozygous configuration (A) and compound heterozygous configuration with a severe variant (B).

