## Supplemental Methods for "Compendium of clinical variant classification for 2,247 unique *ABCA4* variants to improve genetic medicine access for Stargardt Disease"

For each ACMG/AMP classification step the specific application to *ABCA4* data is described below. The steps "PS1 Same amino acid change", "PM3 BP2 Cis/trans testing", "PM5 Novel missense at the same position" and "PS3 BS3 Functional studies" were executed at the end and repeated as described below to increase the number of variant classifications in the categories pathogenic, likely pathogenic, likely benign and benign.

The classification steps "PM1 Mutational hot spot and/or critical and well-established functional domain”, "PP2 BP1 Variant spectrum", "PP4 Using phenotype to support variant claims" as well as the criterium "BS2 Use of control populations" were not applied in this study since these steps are not applicable for *ABCA4*. “PP1 BS4 Segregation analysis” was not used in this study because these data were not consistently available.

*PVS1 Null Variants*

Null variants were categorized as described by Abou Tayoun and others (Abou Tayoun, Pesaran et al. 2018) with some exceptions as described below.

For canonical splice site variants, it is indicated to estimate the impact of the variant on the open reading frame. Where possible, this was based on experimental data. If those data were not available the SpliceAI prediction software was used, as it was proven to be the most accurate for *ABCA4* predictions for non-canonical splice site variants [1] and as it is able to incorporate a wider context of the gene compared to most other splice prediction tools [2].

The PVS1 classification was allocated to all canonical splice site variants that were in the direct vicinity (at a maximum of ten nucleotides away) of non-canonical splice site variants that were experimentally proven to lead to full exon skipping of an exon that did not consist of a multiple of three nucleotides (a so-called ‘out-of-frame exon’) and/or alternative splicing defects that lead to the disruption of the reading frame in midigene assays [3], based on the assumption that if a non-canonical splice site change disrupts the splice site completely, a canonical splice site change will most likely also fully disrupt the splice site.

Canonical splice sites for which no such experimental data were available, were annotated based on SpliceAI predictions. Based on these predictions, the possibility of in-frame and out-of-frame exon skipping as well as alternative splicing was considered. In short, SpliceAI scores were used to predict the effect of the variants on new splice site creation as well as canonical splice site loss. Small indels, which cannot get a score in SpliceAI, were annotated with CI-SpliceAI scores ([VCF Submission - CI-SpliceAI](https://ci-spliceai.com/)), computed with the online CI-SpliceAI tool. A delta score cut-off of ≥0.2 was used to estimate a true predicted effect, which is slightly more conservative than the cut-off calculated by Riepe et al [1]. Exon skipping was only considered to be true if both splice site variants were predicted to be lost. Exon elongation or shortening was considered to be true only if both a splice site was lost and a new splice site was predicted. Variants that were predicted to lead to out-of-frame exon skipping, out-of-frame exon elongation, out-of-frame exon shortening, or would lead to elongation creating a stop codon, were given the PVS1 classification. Variants that were predicted to lead to the loss of only one splice site without having a new splice site predicted, variants where two splice sites were lost but only one alternative splice site was predicted and variants predicted to lead to in-frame exon skipping, were given the ‘PVS1_Strong’ classification based on the assumption that all exons in *ABCA4* are critical to protein function. Variants that were predicted to lead to an in-frame exon elongation/shortening without creating a new stop codon were given the ‘PVS1_Moderate’ classification. Finally, canonical variants at the acceptor splice site of the last exon were annotated with ‘PVS1_Moderate’ based on the prediction of an introduction of 26 random amino acids from the 3’ untranslated region.

Finally, large in-frame deletions (>50% of the gene) that included the loss of the start codon, were annotated with PVS1.

*PM6 De novo variants*

Reported *de novo* variants (n=3) were classified with the PM6 criterium according to the recommendation [svi_proposal_for_de_novo_criteria_v1_1.pdf (clinicalgenome.org)](https://clinicalgenome.org/site/assets/files/3461/svi_proposal_for_de_novo_criteria_v1_1.pdf). Since it was impossible to check the identity of the parents of affected individuals with *de novo* variants, PM6 was applied instead of PS2. According to the recommendations, 0.5 points were attributed to each *de novo* variant with unconfirmed parental relationships, based on the phenotype consistency category “Phenotype is consistent with the gene but not highly specific”. The final evidence strength was based on the total amount of points per variant and Table 2 of the aforementioned recommendation. Of note, two reported *de novo* variants, c.3106G>A and c.4217A>G, were excluded. These were reported in the same individual, which seems unlikely since the variant c.983A>T was found in this individual as well, which is known to occur as a complex allele with c.3106G>A, i.e. c.[983A>T;3106G>A]. This raises the possibility of a sample error [4].

*Variant Frequency and Use of Control Populations (PS4 PM2_Supporting BS1)*

Due to known reduced penetrance in *ABCA4*, BA1, which is the stand alone criterium of benign impact for variants with an allele frequency >0.05, was not used in this study and the frequent variant c.5603A>T was submitted to the Exception List Nomination Form for a BA1 modification as suggested elsewhere [5]. For BS1 a cut-off allele frequency of 0.0163 was used based on the Whiffin/Ware calculator [Frequency Filter (cardiodb.org)](http://cardiodb.org/allelefrequencyapp/) with the conservative settings of a prevalence of 1:7,500, allelic and genetic heterogeneity of 1 and penetrance of 50%. Variants that met this cut-off in any of the gnomAD populations were assigned the BS1 category. The PM2 criterium indicates that if a variant is absent or extremely rare in a control population, there is evidence for this variant to be pathogenic. The evidence is decreased to ‘PM2_Supporting’ according to [pm2_-_svi_recommendation_-_approved_sept2020.pdf (clinicalgenome.org)](https://clinicalgenome.org/site/assets/files/5182/pm2_-_svi_recommendation_-_approved_sept2020.pdf). ‘PM2_Supporting’ was applied to variants with an allele frequency <0.0001 in each of the gnomAD populations, based on the observation that ~95% of the variants categorized as (likely) pathogenic in Cornelis et al., 2017 [6, 7] have an allele frequency of <0.0001 and no variants categorized as (likely) benign had an allele frequency <0.0001. ‘PM2_Supporting’ was only assigned if PS4 was not already assigned.

Before performing an allele frequency analysis, we compared the entries of affected individuals to one another to identify possible duplicates. Cases having the same ≥2 variants, that showed to have a different gender, different age at onset or different ethnicity, were determined to be unique individuals. Whenever the remaining possible duplicates were published by at least one overlapping author, entries were removed from the dataset.

For the allele frequency analysis, we then included all individuals with likely biallelic variants that were not homozygous consanguineous cases. This list included all individuals in which two or more variants were reported. Thereafter, data from individuals were excluded from the dataset when they had exactly two variants identified, without segregation analysis being performed and these variants were also found as part of a complex allele at least twice in the dataset (i.e. c.[1A>G;6089G>A], c.[455G>A;3322C>T], c.[455G>A;6320G>A], c.[769-784C>T;5603A>T], c.[983A>T;3106G>A], c.[1015T>G;5603A>T], c.[1622T>C;3113C>T], c.[1622T>C;4328G>A], c.[2570T>C;5882G>A], c.[2588G>C;5603A>T], c.[3210_3211dup;5603A>T], c.[3322C>T;6320G>A], c.[3386G>T;6718A>G], c.[3758C>T;5882G>A], c.[4222T>C;4918C>T], c.[4253+43G>A;5603A>T], c.[4469G>A;5603A>T], c.[4918C>T;5882G>A], c.[4926C>G;5044_5058del], c.[5282C>G;6316C>T], c.[5329A>T;5603A>T], c.[5461-10T>C;5603A>T]) as well as for the known c.[769-784C>T;5882G>A] allele [8]. Mono-allelic cases were excluded for the two reasons that they might not be *ABCA4*-retinopathy cases and that they likely contain duplicate entries. Finally, 127 alleles from 127 homozygous consanguineous cases were added to the data. Only one allele per individual was included as both alleles most likely have the same origin and therefore represent the same allele in the population.

The variant frequency of the biallelic ABCA4-retinopathy individuals was compared to the earlier published biallelic person (BAP) based genetic ancestry matched (GAM) gnomAD control group (Cornelis et al., 2022) and was tested with a one-tailed Fisher’s Exact test. P-values were corrected with the False Detection Rate from Benjamini and Hochberg 1995 [9]. In short, the BAP gnomAD control group is a control group that endeavors to match the genetic ancestry of published biallelic *ABCA4­*-retinopathy affected individuals based on reported genetic ancestry and estimated genetic ancestry. This control dataset was based on gnomAD version 2.1.1. To determine the allele count in the BAP gnomAD control group, we multiplied the calculated BAP allele frequency with the calculated allele number. For variant locations absent in gnomAD we used the median Allele Number of the BAP matched gnomAD dataset of either exonic and near exonic variants from biallelic *ABCA4*-retinopathy individuals, near exonic being <100 nucleotides away from the exon, or intronic to estimate the allele number on the genomic position of the variant. For structural variants, insertions and deletions >49 base pairs, we used the median of structural variants in gnomAD, since population data were not downloadable.

*PM4 Large In-Frame Deletions Insertions In Conserved Areas*

The PM4 category was assigned to variants leading to a stop loss, variants affecting more than one amino acid or variants affecting one amino acid in which two nucleotides had a PhyloP of 7.367 or higher, which was based on the PhyloP Supporting *in silico* strength from Pejaver et al., 2022 [10].

*PP3 BP4 Computational In Silico Data*

Although Richards et al. suggest to only assign PP3 when all used *in silico* predictions indicate pathogenicity [11], it was decided to perform the *in silico* analysis for splicing and other predictions in parallel. The highest resulting pathogenic criterium was assigned. SpliceAI was used to predict splicing effects. A maximal distance of 2,000 nucleotides between variant and effect was used and only the strongest prediction was considered. For insertion deletion variants, which cannot get a score from SpliceAI, the online tool CI-SpliceAI was used ([VCF Submission - CI-SpliceAI](https://ci-spliceai.com/)). A cut-off of ≥0.2 was used to determine whether a prediction was considered a likely true effect and lead to assignment of PP3. Finally, PP3 was not assigned to canonical splice site variants, since those were already assigned a PVS1 criterium based on SpliceAI predictions unless the predictions were overruled by experimental data as described above.

In parallel, REVEL was used for missense variants and CADD for other variants affecting up to 50 bp. Variants >50 bp did not undergo an *in silico* prediction. Based on Pejaver et al., 2022 [10], missense variants with a REVEL score ≥0.644 and <0.773 were assigned PP3 and variants with a REVEL score ≥0.773 were assigned ‘PP3_Moderate’. Similarly, remaining variants with a CADD score ≥25.3 and <28.1 were assigned PP3 and variants with a CADD score ≥28.1 were assigned ‘PP3_Moderate’. For the assignment of BP4, missense variants with a REVEL score >0.183 and ≤0.290 were assigned BP4 and variants with a REVEL score ≤0.183 were assigned ‘BP4_Moderate’. Other variants with a CADD score >17.3 and ≤20 were assigned BP4 and variants with a CADD score ≤17.3 were assigned ‘BP4_Moderate’. For the BP4 category of CADD an upper cut-off of 20 was applied, a widely used cut-off for pathogenicity.

### For *in silico* programs requiring a vcf input file, [GitHub - counsyl/hgvs: HGVS variant name parsing and generation](https://github.com/counsyl/hgvs) was used (downloaded in August 2021). Duplication variants were manually adjusted when necessary.

*BP7 Synonymous Variants*

Synonymous variants that are unlikely to create splicing defects were assigned BP7. Many *ABCA4* variants located at the outer sides of exons are known to create splice defects. Therefore, all synonymous variants located at the first three or last three nucleotides of an exon were excluded from the BP7 criterium. Furthermore, synonymous variants with a SpliceAI prediction of 0.19 or higher were also excluded from being assigned BP7 based on Riepe et al [1].

**Iterated steps**

The following categories depend on the classification of other variants and were therefore executed at the end, some with several iterations.

*PM3 BP2 Cis/trans* *Testing*

The PM3 category was performed first according to the recommendation: [svi_proposal_for_pm3_criterion_-_version_1.pdf (clinicalgenome.org)](https://www.clinicalgenome.org/site/assets/files/3717/svi_proposal_for_pm3_criterion_-_version_1.pdf). In short, variants occurring in likely biallelic individuals were assessed. Depending on the pathogenicity of the variant a variant of interest occurs with, the variant of interest gets a score between 0.0 and 1.0, depending on phasing data. For example, if a variant of interest is confirmed to be *in trans* with a known pathogenic null variant, then the variant of interest gets 1.0 point for this occurrence. To avoid wrongly assigning a pathogenic criterium to a variant of interest occurring *in* *trans* with a pathogenic variant by chance due to the high frequency of the latter, the variant of interest did not get any points assigned to it if the pathogenic variant *in trans* did not meet the PM2 criterium - occurring with an allele frequency less than 0.0001. For variants that occurred with an allele frequency of >0.0001 themselves we applied the following correction: the number of points assigned to those variants was multiplied with 0.0001 divided by the allele frequency of the variant. The complex alleles c.[1622T>C;3113C>T] and c.[5461-10T>C;5603A>T] were added to the list of pathogenic variants since they are well-known and frequently occurring penetrant pathogenic alleles.

The BP2 criterium was subsequently applied to variants that occurred with a rare (PM2) (likely) pathogenic variant *in* *cis* and that was not reported to occur as a single variant based on phasing data.

*PS3 BS3 Functional Studies*

The PS3 and BS3 criteria were assigned based on the recommendations of Brnich et al. [12]. It was decided that the disease mechanism for STGD1 is understood enough and that midigene assays as well as protein assays model the disease well enough for use of this criterium. These assays contained both wild-type (WT) cDNA constructs and proteins, and cDNA constructs and proteins having known pathogenic variants. Since the assays have been broadly accepted historically, the use of multiple replicates was not necessary.

First, functional data from midigene assays were assessed according to Brnich et al. (Figure 1): looking at both the highest amount of residual WT RNA of non-missense likely pathogenic and pathogenic classified variants and the lowest amount of WT RNA of likely benign and benign classified variants. However, since there was an overlap between (likely) benign and (likely) pathogenic variants in terms of produced WT RNA in the range of 61-76% of WT RNA, we decided on assigning non-missense variants with >80% WT RNA ‘BS3_Supporting’, variants with ≥20% and <50% WT RNA ‘PS3_Supporting’, and variants with <20% WT RNA ‘PS3_Moderate’. The ‘PS3_Moderate’ category is higher than advised by Brnich et al., but it was decided that for these functional assays <20% WT RNA is strong enough evidence to reach Moderate. The variant c.4539+2028C>T, which shows 85% WT RNA expression in photoreceptor progenitor cells [13], was excluded from this list since genotype-phenotype correlations indicate that it is pathogenic. As observed for a few variants that result in a ‘retina-specific’ or ‘retina-enhanced’ splice defect, photoreceptor progenitor cells and retinal organoids do not always show the complete splice defect that is present in the proband’s retinae [13, 14].

Known (likely) pathogenic missense variants that have an F-index as published by Curtis et al., were used to determine an F-index cut-off value for assigning PS3 to missense variants [15]. It was attempted to assess the data as indicated by Brnich et al., but similarly to the midigene assay data an overlap is observed between pathogenic variants and the WT F-index value. However, since the log10 and square root transformed average age at onset of individuals with either homozygous variants or compound heterozygous variants where one variant is known to be severe, correlated really well with the F-index of those variants (Pearson correlations of r(22)=0.6807, p = .000251) and r(40)=0.5269, p = .000337) respectively, Figure S2), it was decided to assign ‘PS3_Moderate’ to variants with an F-index <0.15 and ‘PS3_Supporting’ to variants with an F-index ≥0.15 and <0.50. Log10 and square root transformations were applied to obtain a normal distribution of the data, allowing to calculate the Pearson correlation.

The following steps PS1 and PM5 were executed once, after all the aforementioned steps, and were not iterated to prevent circular reasoning.

*Same amino acid change (PS1)*

Variants that caused the same amino acid change as a known likely pathogenic or pathogenic variant that is not known to cause a splicing effect were assigned PS1.

*Novel missense at the same position (PM5)*

Variants that caused a different amino acid change at the same position as another missense change that was assigned likely pathogenic or pathogenic that is not known to cause a splicing effect were assigned PM5.

After the execution of PS1 and PM5, the step PM3 BP2 *Cis/trans* Testing was executed in iteration until no further changes were present. Since the cut-off values for the PS3 BS3 Functional Studies were already defined, this step was not iterated.
