## Supplementary figures and images for "Compendium of clinical variant classification for 2,247 unique *ABCA4* variants to improve genetic medicine access for Stargardt Disease"

### Figure S1

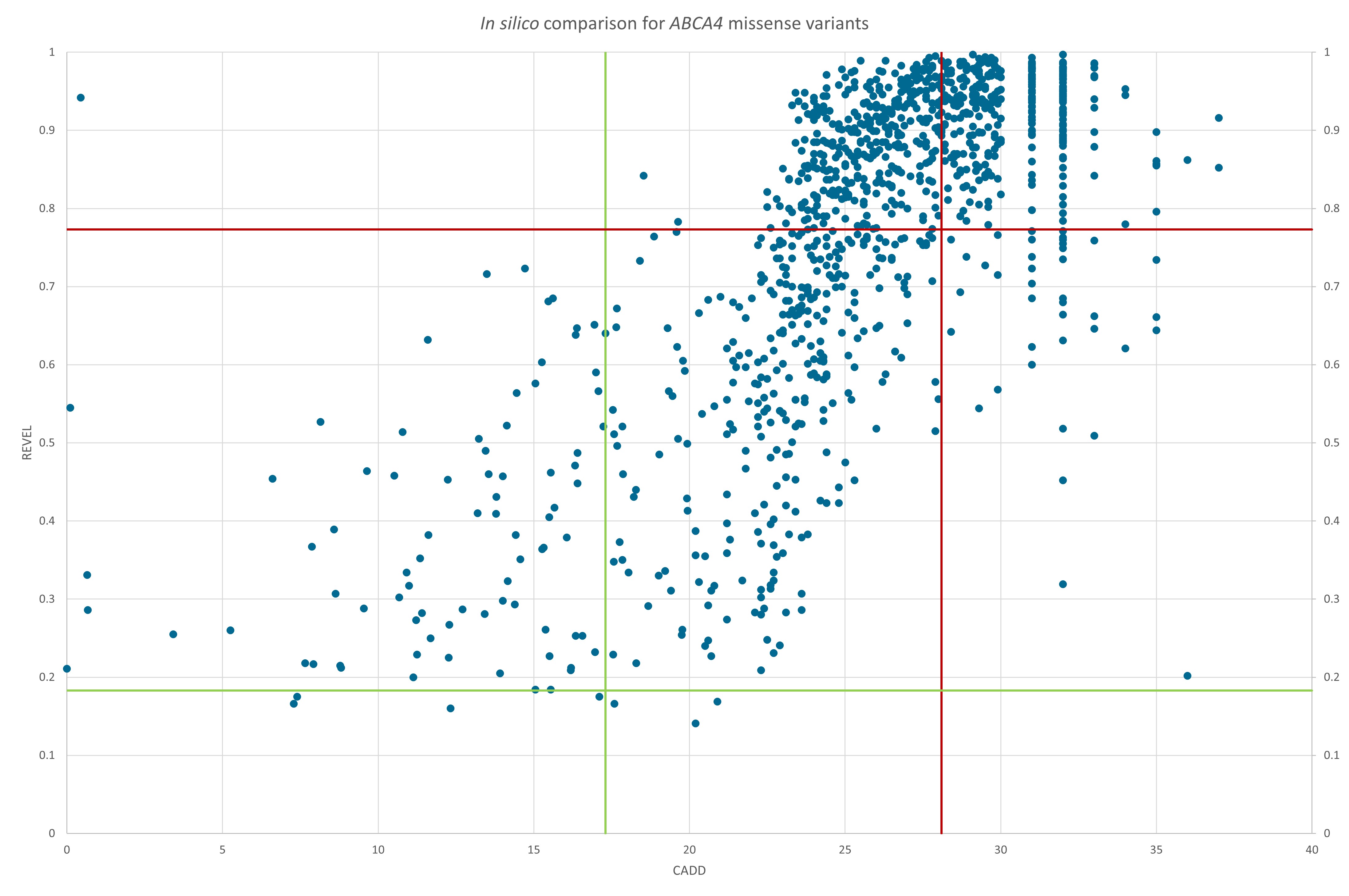

### Figure S2

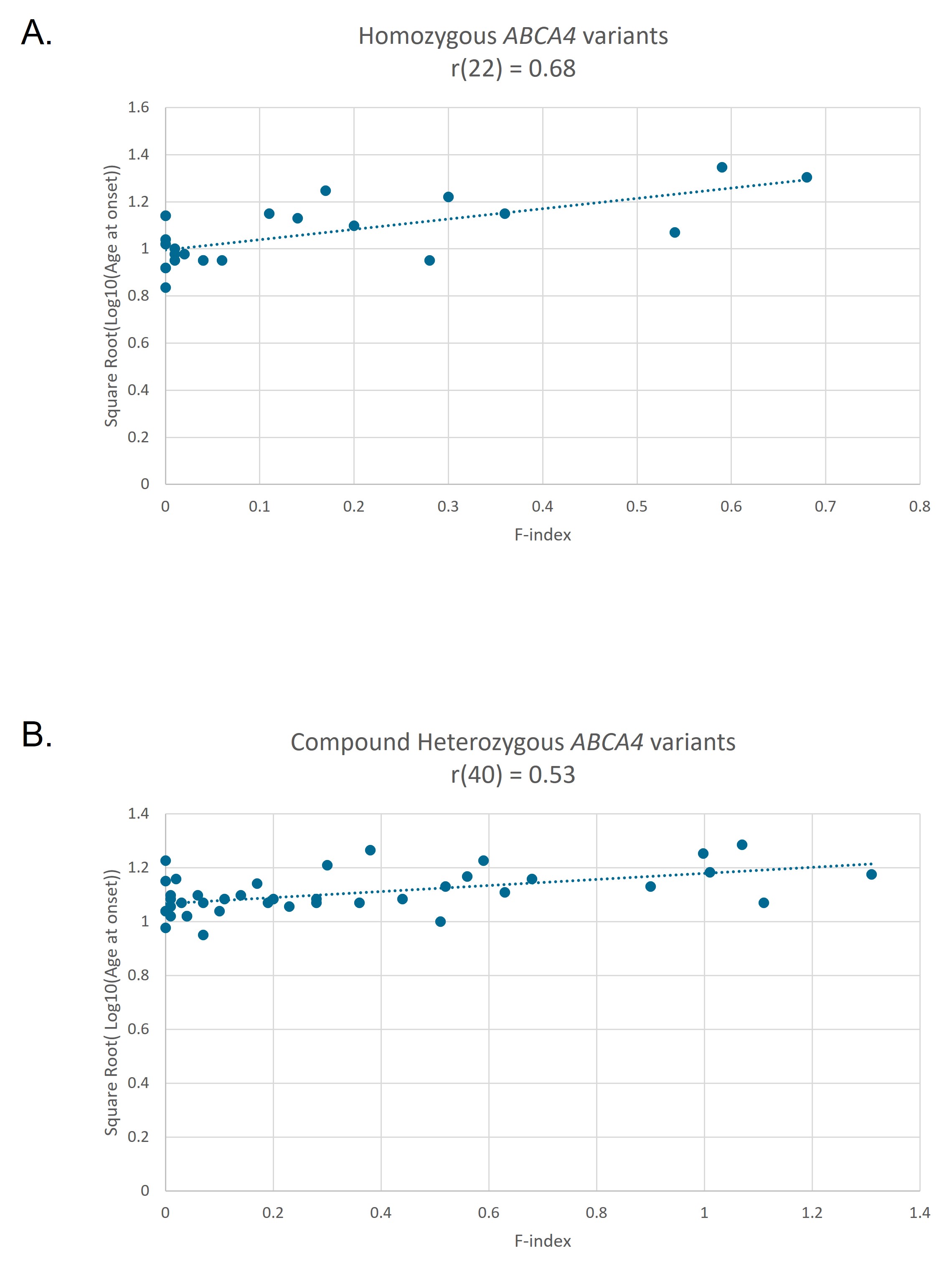
