## Supplementary material for "Compendium of clinical variant classification for 2,247 unique *ABCA4* variants to improve genetic medicine access for Stargardt Disease": Table S11 - Frequent pathogenic variants

Table S11. Three most frequent (Likely) Pathogenic variants per gnomAD population

| **gnomAD population** | **Allele frequency** | **DNA variant** | **Protein variant** | **ACMG/AMP classification** |
| --- | --- | --- | --- | --- |
| **African** | 0.0073 | c.2971G>C | p.(Gly991Arg) | Pathogenic |
|  | 0.0044 | c.2791G>A | p.(Val931Met) | Likely Pathogenic |
|  | 0.0030 | c.2966T>C | p.(Val989Ala) | Pathogenic |
| **Latino/ Admixed American** | 0.0019 | c.5882G>A | p.(Gly1961Glu) | Pathogenic |
|  | 0.0017 | c.3386G>T | p.(Arg1129Leu) | Pathogenic |
|  | 0.0015 | c.2453G>A | p.(Gly818Glu) | Pathogenic |
| **Ashkenazi Jewish** | 0.0233 | c.5882G>A | p.(Gly1961Glu) | Pathogenic |
|  | 0.0023 | c.4139C>T | p.(Pro1380Leu) | Pathogenic |
|  | 0.0021 | c.4594G>A | p.(Asp1532Asn) | Pathogenic |
| **East Asian** | 0.0020 | c.6119G>A | p.(Arg2040Gln) | Pathogenic |
|  | 0.0019 | c.1531C>T | p.(Arg511Cys) | Likely Pathogenic |
|  | 0.0016 | c.71G>A | p.(Arg24His) | Pathogenic |
| **Finnish European** | 0.0056 | c.3113C>T | p.(Ala1038Val) | Likely Pathogenic |
|  | 0.0011 | c.2588G>C | p.[Gly863Ala,Gly863del] | Pathogenic |
|  | 0.0007 | c.1622T>C | p.(Leu541Pro) | Pathogenic |
| **non-Finnish European** | 0.0078 | c.2588G>C | p.[Gly863Ala,Gly863del] | Pathogenic |
|  | 0.0038 | c.5882G>A | p.(Gly1961Glu) | Pathogenic |
|  | 0.0023 | c.3113C>T | p.(Ala1038Val) | Likely Pathogenic |
| **South Asian** | 0.0138 | c.5882G>A | p.(Gly1961Glu) | Pathogenic |
|  | 0.0020 | c.2588G>C | p.[Gly863Ala,Gly863del] | Pathogenic |
|  | 0.0007 | c.859-9T>C | p.[=,Phe287_Arg452del] | Pathogenic |
| **Other** | 0.0057 | c.5882G>A | p.(Gly1961Glu) | Pathogenic |
|  | 0.0036 | c.2588G>C | p.[Gly863Ala,Gly863del] | Pathogenic |
|  | 0.0022 | c.3113C>T | p.(Ala1038Val) | Likely Pathogenic |
