## Supplementary material for "Compendium of clinical variant classification for 2,247 unique *ABCA4* variants to improve genetic medicine access for Stargardt Disease": Table S12 - Previously published frequent pathogenic variants

Table S12 Previously reported frequent Pathogenic variants based on literature

| **DNA variant** | **Protein variant** | **ACMG/AMP classification** | **gnomAD allele frequency (population with highest frequency)** | **Region or population (reference)** |
| --- | --- | --- | --- | --- |
| c.768G>T | p.Leu257Valfs*17 | Pathogenic | 0.000186  (non-Finnish European) | Netherlands, [1] |
| c.859-9T>C | p.[=,Phe287_Arg452del] | Pathogenic | 0.000686  (South Asian) | American of South Asian descent, [2, 3] |
| c.[1622T>C;3113C>T] | p.[(Leu541Pro);(Ala1038Val)] | Likely Pathogenic | Not applicable | Germany and Poland, [4-7] |
| c.2588G>C | p.[Gly863Ala,Gly863del] | Pathogenic | 0.00784  (Non-Finnish European) | Western Europe, [8, 9] |
| c.2894A>G | p.(Asn965Ser) | Pathogenic | 0.000434924  (East Asian) | Denmark and China, [10, 11] |
| c.3386G>T | p.(Arg1129Leu) | Pathogenic | 0.001665  (Latino/Admixed American) | Spain, [12] |
| c.4139C>T | p.(Pro1380Leu) | Pathogenic | 0.002318393  (Ashkenazi Jewish) | Ashkenazi Jewish, [13] |
| c.4254-37_4254-15del | p.(?) | Not categorized | absent | Ashkenazi Jews, [14] |
| c.4469G>A | p.(Cys1490Tyr) | Pathogenic | 0.00013  (non-Finnish European) | South African of European descent, [15] |
| c.4539+2001G>A | p.[=,Arg1514Leufs*36] | Pathogenic | 6.48E-05  (Non-Finnish European) | Belgium, [16-18] |
| c.5318C>T | p.(Ala1773Val) | Pathogenic | 0.000463  (Latino/Admixed American) | Mexico, [19, 20] |
| c.5882G>A | p.(Gly1961Glu) | Pathogenic | 0.023341049  (Ashkenazi Jewish) | Somalia and American of South Asian descent, [2, 21, 22] |
| c.5917del | p.(Val1973*) | Pathogenic | 0.000163  (South Asian) | Hungary and Bulgaria, [23, 24] |
| c.6320G>A | p.(Arg2107His) | Likely Pathogenic | 0.020794935  (African) | African American, [25] |
